# Ultrarare Variants in DNA Damage Repair Genes in Pediatric Acute-Onset Neuropsychiatric Syndrome or Acute Behavioral Regression in Neurodevelopmental Disorders

**DOI:** 10.1101/2024.02.20.24302984

**Authors:** Janet L. Cunningham, Jennifer Frankovich, Robert A. Dubin, Erika Pedrosa, Refıa Nur Baykara, Noelle Cathleen Schlenk, Shahina B. Maqbool, Hedwig Dolstra, Jacqueline Marino, Jacob Edinger, Julia M. Shea, Gonzalo Laje, Sigrid M.A. Swagemakers, Siamala Sinnadurai, Peter J. van der Spek, Herbert M. Lachman

## Abstract

Acute onset of severe psychiatric symptoms or regression may occur in children with premorbid neurodevelopmental disorders, although typically developing children can also be affected. Infections or other stressors are likely triggers. The underlying causes are unclear, but a current hypothesis suggests the convergence of genes that influence neuronal and immunological function. We previously identified 11 genes in Pediatric Acute-Onset Neuropsychiatry Syndrome (PANS), in which two classes of genes related to either synaptic function or the immune system were found. Among the latter, three affect the DNA damage response (DDR): *PPM1D, CHK2,* and *RAG1*. We now report an additional 17 cases with mutations in *PPM1D* and other DDR genes in patients with acute onset of psychiatric symptoms and/or regression that were classified by their clinicians as PANS or another inflammatory brain condition. The genes include clusters affecting p53 DNA repair (*PPM1D*, *ATM, ATR*, *53BP1,* and *RMRP*), and the Fanconi Anemia Complex (*FANCE, SLX4/FANCP, FANCA, FANCI,* and *FANCC*). We hypothesize that defects in DNA repair genes, in the context of infection or other stressors, could lead to an increase in cytosolic DNA in immune cells triggering DNA sensors, such as cGAS-STING and AIM2 inflammasomes. These findings could lead to new treatment strategies.

## Introduction

We recently reported the first whole exome and whole genome sequencing (WES and WGS, respectively) analysis in Pediatric Acute-Onset Neuropsychiatry Syndrome (PANS) in which ultrarare variants were found in 11 genes in 21 cases^1^. The genes harboring ultrarare variants (minor allele frequency <0.001) showed extensive heterogeneity but clustered to those that affect innate and adaptive immunity; *PPM1D* (3 cases), *NLRC4* (4 cases), *RAG1* (3 cases), *PLCG2,* and *CHK2*, or genes that affect synaptic function; *SGCE* (2 cases), *CACNA1B* (2 cases), *SHANK3* (3 cases), and one case each for *GRIN2A, GABRG2,* and *SYNGAP1.* Of the 21 subjects, five were diagnosed with autism spectrum disorder (ASD) before their deterioration that met PANS criteria. In addition to ASD, four cases were diagnosed with another neurodevelopmental disorder (NDD), one of whom had Jansen de Vries Syndrome (JdVS), which is caused by truncating mutations in *PPM1D* exons 5 or 6 that cause a gain-of-function effect by suppressing p53 and other proteins involved in the DNA damage repair response (DDR) (e.g., MDM2, ATM, CHK1, CHK2, ATR, and H2AX)^2–6^. The high prevalence of PANS superimposed on pre-existing NDD is consistent with the observations of other groups^7, 8^.

PANS is thought to be an inflammatory disorder that primarily affects the deep brain grey matter (primarily the basal ganglia but the thalamus and amygdala may be involved in more extensive cases). This hypothesis is based on the cardinal symptoms of PANS and is supported by four neuroimaging studies showing the following: swelling of the basal ganglia during the acute stage, microglial activation in the caudate and globus pallidus, grey/white matter differences in the basal ganglia, and microstructural changes throughout the brain but most prominently in the basal ganglia^9–12^.

The clinical diagnosis PANS has core symptoms of abrupt onset obsessive-compulsive disorder (OCD) and/or restricted eating (food aversion), with two or more secondary symptoms: anxiety, emotional lability, irritability, rage, cognitive regression, sleep disturbance, sensory dysregulation, movement abnormalities, urinary symptoms including new onset dysuria or urinary frequency^13^. While the criteria require 2 secondary symptoms, most patients have 5 to 6 secondary symptoms that start abruptly alongside the OCD and/or eating restriction. Autonomic instability (POTS, dilated pupils, increased urinary frequency, enuresis) and hypermobility have also been reported as co-morbid with PANS^13–18^. A deterioration in school performance occurs, exemplified by the loss of previously learned skills and/or new onset procedural learning challenges presumably relating to the basal ganglia inflammation PANS^13–18^. Activities of daily living, extracurricular activities, and social interactions are severely affected during relapses. PANS can occur in children with or without premorbid ASD or other NDD.

For parents, the abrupt onset is a telling feature with a rapid decline in behavior and functioning that is sometimes, literally, overnight or emerging over a period of a week. The course is typically relapsing and remitting, but not all relapses are hyperacute/acute. Relapses are frequently associated with an infection and data in support of a neuroinflammatory etiology for PANS is accumulating. In the subgroup of cases following Streptococcus infections, a range of group A streptococcal (GAS) autoantibodies and antibodies to striatal cholinergic interneurons can be detected^15, 19–21^. Recently, an increased prevalence of folate receptor alpha autoantibodies associated with cerebral folate deficiency and ASD were detected in PANS^22^.

Additionally, cross-sectional prevalence data show that approximately one-third of patients meeting PANS criteria eventually develop enthesitis-related arthritis (ERA) and/or inflammatory back pain further supporting a role for inflammation in this disorder^18, 23^. Furthermore, inflammatory/autoimmune disorders are prevalent in first-degree family members^8, 24, 25^.

As is true in other rare pediatric rheumatological conditions (including ERA and Sydenham chorea), observational studies in PANS suggest that non-steroidal anti-inflammatory drugs (NSAIDs) and corticosteroids may alleviate symptoms and/or shorten the length of the relapses^26–28^. More potent immunomodulators, such as intravenous immunoglobulin (IVIg) and a B-cell inhibitor Rituximab have mixed results, likely due to the genetic heterogeneity and the difficulty in appropriately powering randomized trials in relapsing/remitting conditions^29, 30^.

An immune-based etiology is also suggested in some cases of ASD and NDD, especially acute and subacute regression, which have some symptoms that overlap with PANS and respond to immunomodulators^31–34^. In addition, a strong family history of autoimmune disorders is associated with an increased risk for ASD^35–37^.

We are interested in identifying genetic factors that underlie the development of PANS and acute behavioral regression using next-generation sequencing. In our original paper, two of the ultrarare variants we found are in genes that affect DDR (*PPM1D* and *CHK2*), and one induces DNA breaks (*RAG1*). *RAG1*, along with *RAG2*, codes for endonucleases that induce DNA double-strand breaks responsible for the somatic recombination that generates immune cell diversity in B- and T-lymphocytes^1, 38, 39^. RAG1/RAG2 also affects NK cell differentiation and plays a role in DNA transposition^40, 41^. Double-stranded breaks induced by the RAG complex are repaired by ATM, a serine/threonine kinase that orchestrates DNA repair at double-strand breaks^39, 42–44^.

Our overarching aim is to identify genetic variations that converge on common biological pathways within the heterogeneous population of children with PANS or acute-onset regression with or without premorbid NDD. Given the findings in our first PANS genetic study, one common pathway is DNA repair. This hypothesis is supported by the findings in this paper in which many other genes involved in DNA repair were identified that may contribute to genetic vulnerability.

## Subjects and Methods

### Ethical considerations

This study was conducted in accordance with the Declaration of Helsinki. Parents signed informed consent approved by the Albert Einstein College of Medicine IRB (2022-14636).

#### Subjects

The subjects were identified through connections between the senior investigators and a PANS group called EXPAND, a non-profit European advocacy organization for families of children, adolescents, and adults with immune-mediated neuropsychiatric disorders (https://expand.care/), through the Neuroimmune Foundation, a non-profit organization dedicated to neuroimmune and inflammatory brain conditions (https://neuroimmune.org/about/), The Louisa Adelynn Johnson Fund for Complex Disease (https://tlajfundforcomplexdisease.com/), and the Jansen de Vries Syndrome Foundation (https://jansen-devries.org/). Histories were obtained by the participating physicians and confirmed and collated by one of the senior investigators who interviewed every family member (H.M.L). The cases met the criteria for PANS with or without co-morbid ASD or NDD, or individuals with ASD or other NDDs who had a history of rapid-onset behavioral regression.

### Genetic analyses

Whole exome sequencing (WES) data was available for six cases (**Table**). WES was carried out by different companies Courtagen Diagnostics Laboratory, GeneDx, Baylor Genetics, and for case 10, WES was conducted locally in the Genetics Core at the Albert Einstein College of Medicine, Epigenetics Shared Facility. For the remaining cases, data was available from commercially available panels: Invitae (Primary Immunodeficiency Panel), NHS North West (NDD panel). The genetic variants for cases 1 and 4-12 were verified using Sanger sequencing (see **Table** and **Supplementary Methods**). A comprehensive description of the library preparation and analysis of sequencing data for the Einstein sample (case 10) can be found in the **Supplementary Methods** section. Parents were genotyped for variants found in all cases except cases 10, and 13-16.

**Table.**
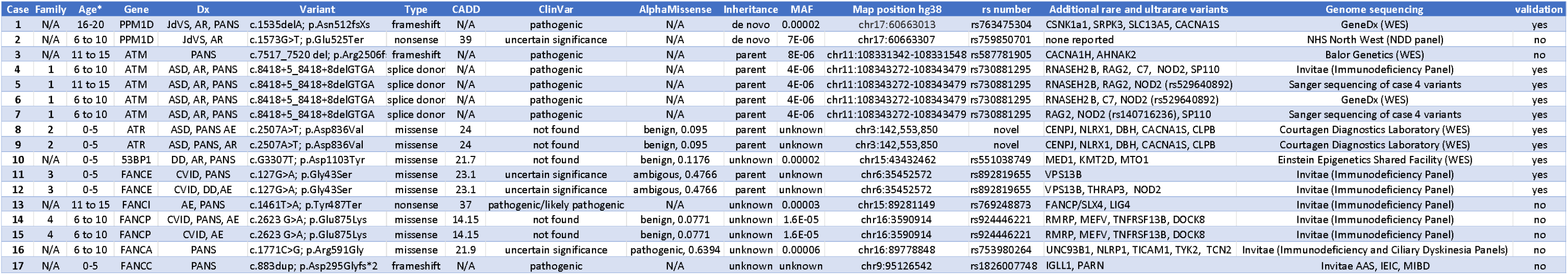
Description of all primary DNA repair variants. *Age range at diagnosis; Dx: diagnosis (JdVS: Jansen de Vries Syndrome, AR: acute regression, PANS: pediatric acute-onset neuropsychiatric syndrome, AE: autoimmune encephalitis, DD: developmental delay, CVID: common variable immune deficiency); variant shows the cDNA and amino acid changes; type shows how the mutation affects gene function; CADD: Combined Annotation score; N/A not available. ClinVar is the predicted pathogenicity based on the ClinVar database; AlphaMissense is the predicted pathogenicity based on amino acid change (see main text); Inheritance pattern shows that the variant in question was either de novo or transmitted from a parent. The sex of the parent was omitted to maintain anonymity; MAF: Minor allele frequency; Map position shows coordinates of variants based on hg38 build; rs number is based on SNP database; Additional variants were found in each case (see Supplemental table for descriptions). Genome sequencing shows the different methods used to detect variants, For case 17, Invitae AAS, IEIC, and MIBD panels are Autoinflammatory and Autoimmunity Syndromes, Inborn errors of immunity and Cytopenia, and Monogenic Inflammatory Bowel Disease, respectively.

### Evaluation of Variants

Minor allele frequencies (MAFs) were obtained from the GnomAD database on the UCSC Genome Browser (https://genome.ucsc.edu/). The ClinVar database, Combined Annotation-Dependent Depletion (CADD), and AlphaMissense were used to predict the pathogenicity of mutations^45–48^. https://www.ncbi.nlm.nih.gov/clinvar/. RNAfold was used to predict the secondary structure of RMRP. A detailed explanation is in the **Supplementary Methods** section.

## Results

Ultrarare variants (MAF <0.001) affecting DNA repair genes were found in 17 cases (11 males and 6 females; specific sex of cases are not included to maintain anonymity) including four families with two or more affected individuals. The identified genes are involved in the p53 repair pathway: *PPM1D* (2 cases), *ATM* (5 cases,), *ATR* (2 cases), and *53BP1* (one case) (**Figure 1**), and the Fanconi Anemia Complex (FC): *FANCE* (2 cases), *FANCP/ SLX4* (2 cases), *FANCI*, *FANCA*, and *FANCC* (one case each) (**Figure 2**; see **Table** for details on all p53 and FC gene variants). The FC pathway repairs DNA interstrand crosslinks and interacts with ATM/ATR/p53 DNA repair pathways **(Figure 2)**^49–54^. In addition, ultrarare variants were found in 34 other genes, many of which have effects on DNA repair and innate immune function (see **Supplementary Table** for details on all additional variants described in this paper). STRING was used to show the connections between the DNA repair genes and other variants found in this study, which centered around ATM and p53 (**Figure 3**). The cases and genetic variants with their potential impact on the phenotypes are described below.

**Figure 1.**
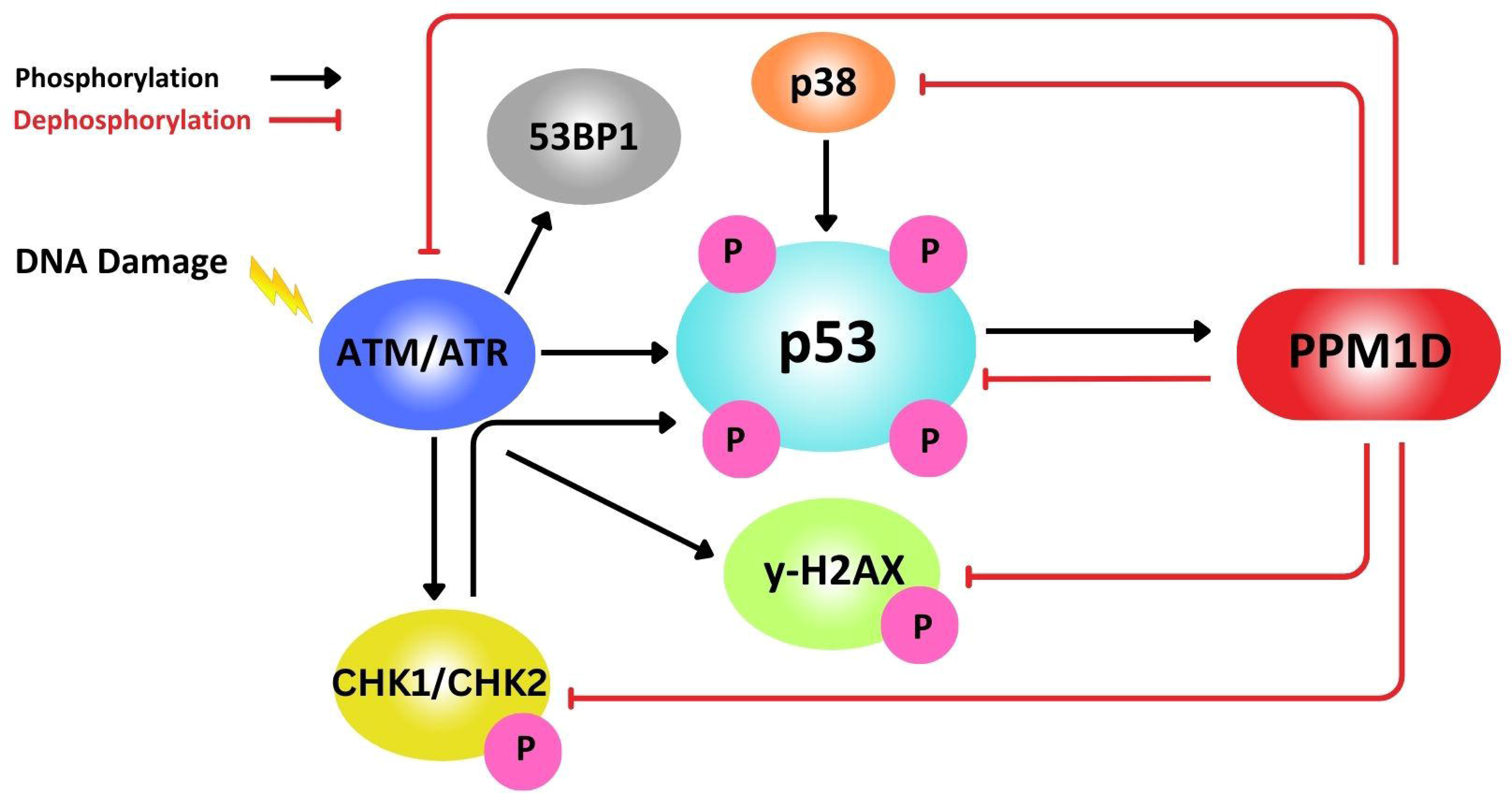
p53 DNA repair pathway. A simplified depiction showing the regulation of p53 tumor suppressor activity by phosphorylation triggered by ATM, ATR, CHK1, and CHK2 kinases, and dephosphorylation by PPM1D. P53 activation can induce apoptosis or cell cycle arrest, which will eliminate DNA-damaged cells or provide an opportunity to repair the damage, respectively. Loss of function mutations in *ATM* or gain of function mutations in *PPM1D*, such as those found in patients with acute neuropsychiatric decompensation in cases 1-7, would be expected to lead to a decrease in p53 activity.

**Figure 2.**
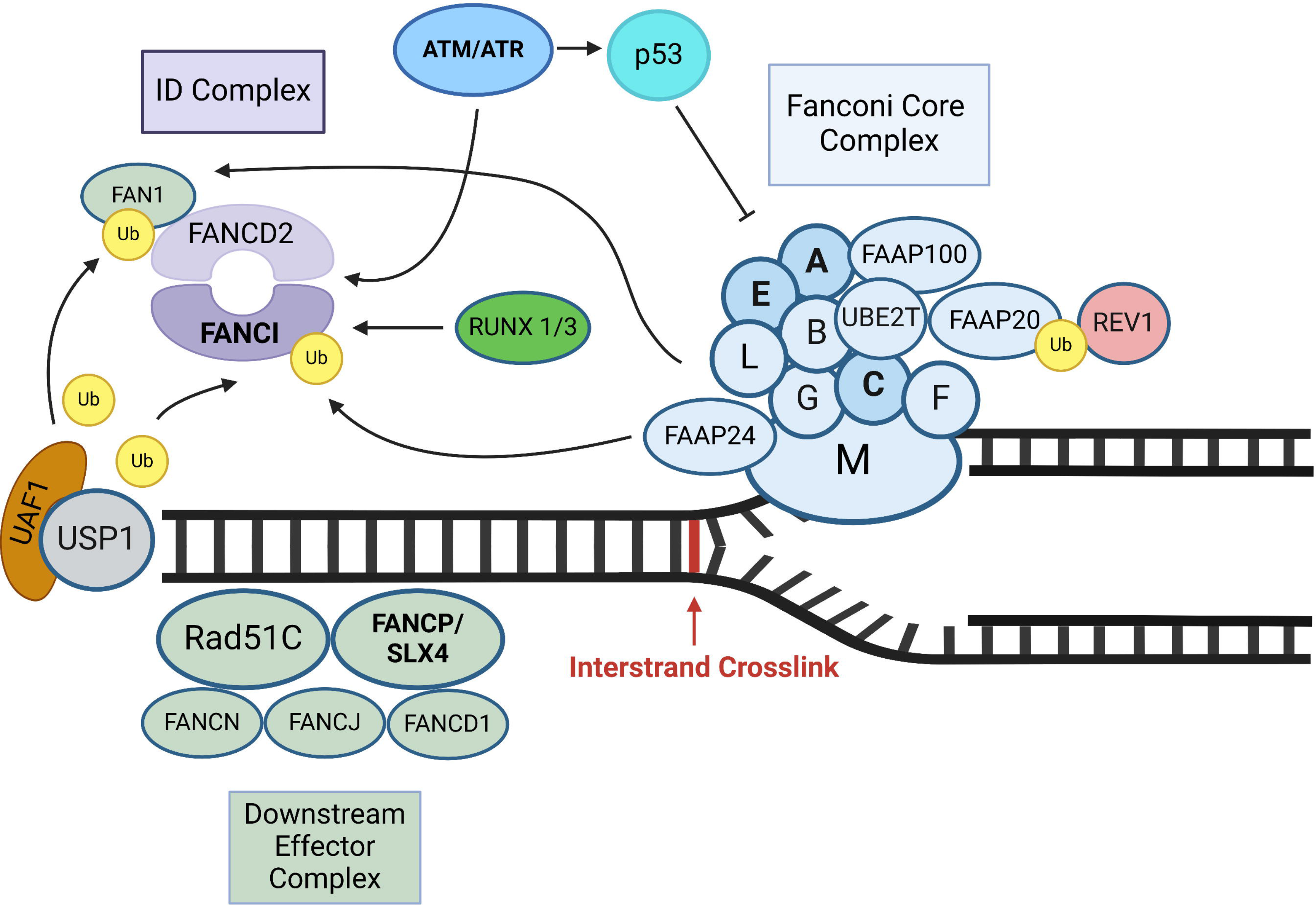
Fanconi Core Complex. The Fanconi Core Complex of proteins repairs interstrand crosslink breaks (Figure adapted from reference 49). The letters are abbreviations for the Fanconi complex proteins (i.e., “A” is FANCA; “E” is FANCE, etc.). Monoubiquitinated (Ub) FANCD2 and FANCI form a dimer that binds to crosslink repair sites, which leads to the recruitment of nucleases that repair the DNA lesion (reference 53). All of the Fanconi complex genes described in this paper are depicted in bold type (FANCE, SLX4, FANCE, and FANCA). Interactions between Fanconi proteins with ATM/ATR are shown. The image was generated using biorender https://www.biorender.com/).

**Figure 3.**
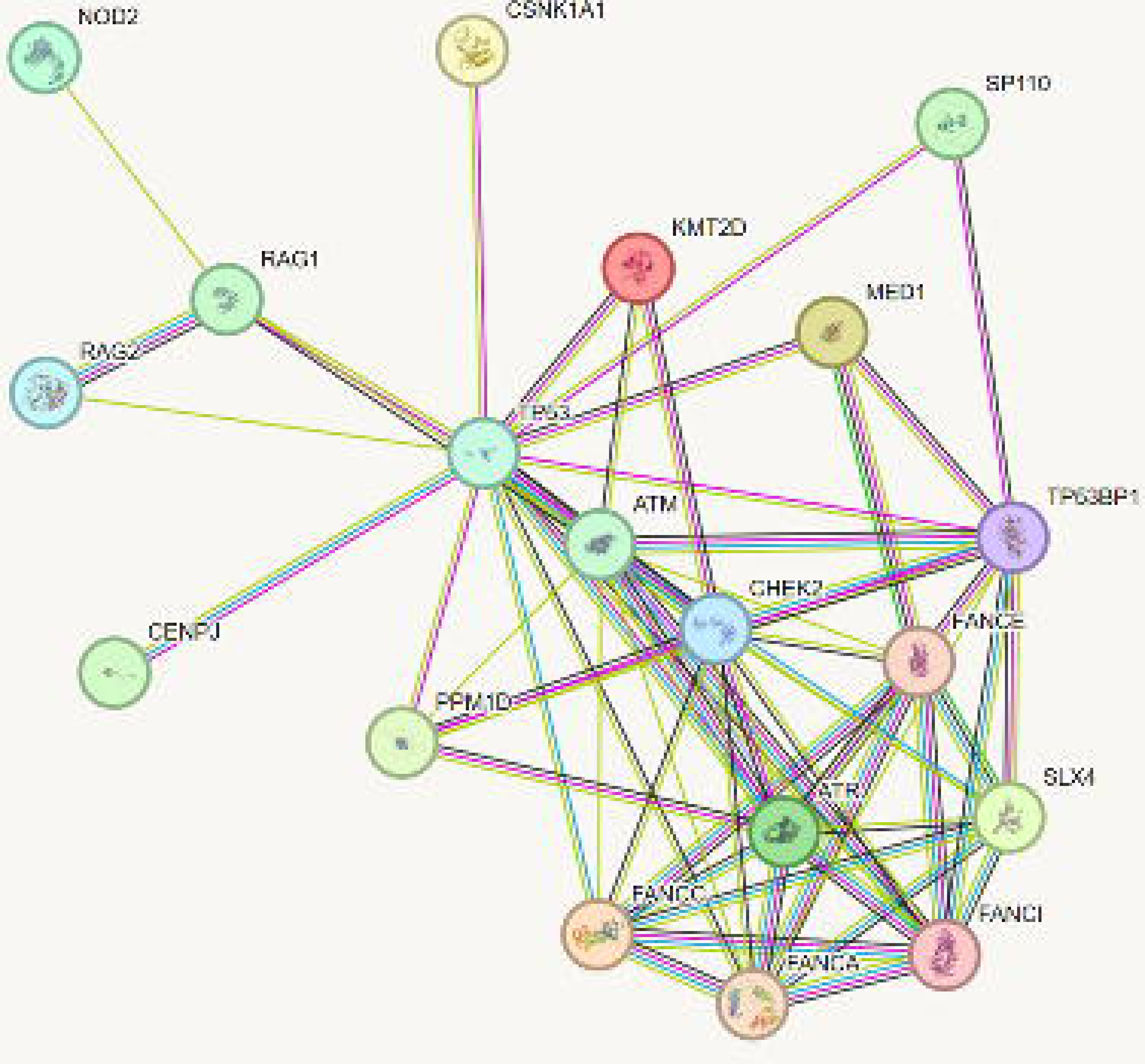
Protein-Protein Interaction Network: STRING. A connectivity network was generated for the candidate genes using IPA software. Central to the network are ATM and p53. Genes described in the paper that do not fit into the connectivity network are not shown (C7, DBH, SLC13AS, SRPK3, NLRX1, MTO1, CACNA1H, CACNA1S, CLPB, AHNAK2). SLX4 = FANCA/SLX4, TP53=p53, and TP53BP1 = 53BP1.

### p53 DNA Repair Pathway (*PPM1D, ATM, ATR, 53BP1*)

### PPM1D (Protein phosphatase, Mg2+/Mn2+dependent 1D)

Case 1 has JdVS with a 1:1 mosaicism for a de novo truncating *PPM1D* variant in exon 6. The patient was diagnosed with mild intellectual disability but was high functioning, requiring minimal support until high school when the patient had an abrupt onset of OCD, anxiety, cognitive regression, insomnia, and psychosis with rapid decline over approximately three weeks. The deterioration coincided with an upper respiratory infection and post-infectious vasculitis. Further regression occurred with the loss of previously mastered global abilities and the eventual development of mutism, which has persisted for several years, unresponsive to psychiatric medications and other clinical and behavioral interventions.

Two additional ultrarare de novo variants were found and are likely pathogenic, one of which is relevant to the DDR hypothesis: a variant in *CSNK1a1,* which codes for Casein Kinase 1 Alpha 1, a p53 inhibitor^55, 56^. Another was found in *SRPK3,* an SRSF Protein Kinase that has no known effect on DDR, although interestingly, expression is inversely correlated with alpha-synuclein levels in mouse models of Parkinson’s disease^57^. Finally, an inherited ultrarare, nonsense mutation was found in the citrate transporter, *SLC13A5* as well as a splice acceptor variant in *CACNA1S*.

Case 2 also has a typical JdVS variant (a truncating mutation in *PPM1D* exon 6) and was diagnosed with ASD and a learning disability. The patient has periods of abrupt onset behavioral regression lasting several days to months, characterized by eating refusal, rage, cognitive deficits, loss of previously learned skills, an inability to walk, and enuresis. The most severe relapses have occurred following infection with SARS-CoV2 and a Group A Streptococcal infection concurrent with shingles.

### ATM (Ataxia-telangiectasia mutated or serine/threonine kinase)

Case 3 carries an ultrarare and pathological frameshift mutation in *ATM*. The patient was typically developing and excelled in school until adolescence when diagnosed with Postural Orthostatic Tachycardia Syndrome (POTS), irritable bowel syndrome gastroparesis, and enthesitis-related arthritis. Soon thereafter, an episode of gastroenteritis led to acute-onset OCD, anxiety, impulsivity, and severe rage; the first of many such episodes. An MRI scan was negative. Relapses responded to IVIg and steroid bursts; however, due to frequent relapses and the development of chronic symptoms, a trial of Rituximab was undertaken, which was helpful. While the patient appeared to respond to immunomodulation, severe symptoms returned when these therapies were tapered or discontinued. Multiple hospitalizations for uncontrolled psychiatric symptoms followed. A parent carries the same ATM variant and has a history of dysautonomia and an inflammatory skin condition.

Other ultrarare variants found in this patient include a de novo variant in *AHNAK2* and compound heterozygosity for two ultrarare *CACNA1H* variants. While there are no obvious connections to DNA repair, AHNAK2 has been implicated in systemic lupus erythematosus (SLE) in two studies and *CACNA1H* mutations are found in ASD and intellectual disabilities^58–61^.

Cases 4-7 are a family with four affected children who all carry a pathogenic parental transmitted ATM splice donor mutation. There is a bilneal parental history of severe autoimmune disorders and hypermobile joints, as well as an extended family history of early-onset cancer, and autoimmune disorders. This is consistent with the increased risk of cancer and autoimmune disorders in individuals with loss-of-function mutations in *ATM* ^62–65^.

Case 4, the proband, was diagnosed with ASD and experienced a typical PANS relapsing-remitting course with abrupt relapses of OCD, food aversion, anxiety, irritability, impulse control problems, oppositional behaviors, tics, and fatigue. Relapses were commonly coincident with infections and were effectively managed by addressing infections with antibiotics and using IVIg. The proband also has a history of Hashimoto’s thyroiditis, joint hypermobility, and consistently low immunoglobulin levels which likely contributed to frequent infections.

Alongside the *ATM* variant, the patient inherited rare and ultrarare variants affecting DNA repair and innate immunity including a likely pathogenic variant in *RNASEH2B*, which codes for Ribonuclease H2, an enzyme that removes ribonucleotides that have been incorporated into replicating DNA, affecting double-stranded break repair kinetics^66^. Homozygosity for this variant leads to Aicardi-Goutières syndrome (AGS), a type I interferonopathy characterized by neurodevelopmental defects and upregulation of type I interferon signaling and neuroinflammation due to the effects of DNA damage^66, 67^. The patient also has ultrarare nonsynonymous variants of unknown significance in *RAG2, NOD2*, and a 24-base pair microduplication in *SP110*. Each of these genes has effects on innate immunity, but RAG2 is the only one with a strong connection to DNA repair, as described in the introduction^1, 38, 39^ ^68^. An *NOD2* variant was transmitted from one of the parents. A second ultrarare variant in *NOD2* was identified in the other parent (see **Supplementary Table**).

Cases 5-7 are siblings to case 4 who were all diagnosed with ASD, dysautonomia, and joint hypermobility. They all have a relapsing-remitting course characterized by abrupt onset OCD, anxiety, emotional lability, increased urinary frequency, oppositional behavior, and reduced social interactions, along with cognitive and behavioral regression. Relapses are typically coincident with infection. Case 5 had an MRI that showed right temporal sclerosis.

Cases 6 and 7 have epilepsy, low levels of IgM and IgA, and relapses improve after clearance of infection and treatment with IVIg and/or a corticosteroid burst. Although they were not analyzed by WES, each was analyzed by Sanger sequencing for the variants found in the proband. Each sibling has inherited the *ATM* variant, and cases 5 and 6, the *RNASEH2B* variant. They have also inherited one of the two *NOD2* variants.

### ATR (Ataxia telangiectasia and rad3-related protein)

Cases 8 and 9 are siblings with regressive ASD and a relapsing-remitting course with relapses commonly coincident with infections. They both have a unique, transmitted ultrarare *ATR* variant of unknown significance. One parent has a history of Hashimoto’s thyroiditis and a family history of SLE, and the other parent has a history of joint hypermobility and childhood seizure disorder. ATR is a serine/threonine kinase and DNA damage sensor that functions as a master regulator of the DDR^69–73^. The proband, case 8, was a typically developing child with hypermobility who was well until abrupt behavioral regression occurred following “croup” at which time a diagnosis of ASD was made. The patient subsequently developed a relapsing-remitting course characterized by abrupt onset OCD, aggression, and mutism. Relapses appeared to be responsive to NSAIDs.

The proband’s sibling, Case 9, was well until the development of an abrupt behavioral regression and loss of previously achieved milestones following a herpesvirus 6 infection, after which a diagnosis of ASD was made. The patient also has a relapsing-remitting course characterized by abrupt onset of OCD, restricted eating, rage, and depression. Relapses are typically coincident with infections. Symptoms typically respond to glucocorticoid bursts. MRI showed atrophy of the hippocampus, amygdala, caudate, and putamen. WES analysis of both cases 8 and 9 also revealed several other ultrarare variants of interest: *NLRX1*, *CENPJ, CACNA1S, DBH, and CLPB.* The most interesting from the DNA repair perspective is a pathogenic frameshift mutation in *CENPJ,* which codes for centromere protein J. Centromere regulation is critical for maintaining genomic stability, and defects lead to aberrant mitosis and activation of the cGAS-STING pathway^73, 74^. It is interesting to note that biallelic mutations in *CENPJ* and *ATR* can both cause Seckel syndrome, which is associated with cell-cycle checkpoint signaling abnormalities^75, 76^. Another variant was found in *NLRX1,* which codes for a key player in the innate immune response that regulates NF-κB expression and has been implicated in autoimmune and inflammatory diseases^77–80^. A third sibling is typically developing and did not inherit the *ATR* variant but carries the *CENPJ, CACNA1S,* and *CLPB* variants.

Thus, the two affected siblings with ASD and acute-onset neuropsychiatric symptoms have ultrarare variants in two genes that affect DNA repair, *ATR,* and *CENPJ,* as well as *NLRX1 DBH, CACNA1S,* and *CLPB*.

### 53BP1 (Tumor suppressor p53-binding protein 1)

Case 10 has a history of developmental delay, and experienced severe regression as a young child characterized by loss of the ability to walk, loss of verbal skills, and new-onset enuresis. Upon WES, we found an ultrarare missense mutation in *53BP1*, a p53-binding protein that regulates DDR ^81–83^. A preceding infection to initial deterioration was not ascertainable.

However, the patient developed a relapsing-remitting course with relapses (abrupt onset of OCD, eating refusal, severe rage, and oppositional behavior) coincident with infections with improvement following treatment infections with antibiotics. The patient also developed Hashimoto’s thyroiditis, scoliosis, joint hypermobility, and gastroparesis. A parent has a history of Raynaud’s and fibromyalgia, and there is a family history of thyroid disease and Parkinson’s Disease. Parents have not been genotyped. In addition to 53BP1, several other ultrarare variants were found that could be playing a role in the clinical state via DNA repair. One is *MED1*, which codes for a mediator complex protein that plays a major role in homologous recombination, base excision repair, the maintenance of genomic stability, and p53-dependant apoptosis^84^. It has also been implicated in SLE^85^. Another is *KMT2D*, which codes for a histone methyltransferase that plays a role in DNA repair and has been implicated in autoimmune disorders^86, 87^. Finally, we found a nonsense mutation in *MTO1*, which regulates oxidative phosphorylation and has been found to affect DNA repair and recombination in yeast ^88, 89^.

### Fanconi Anemia Complex (FANCE, FANCP/SLX4, FANCA, FANCI, and FANCC)

### FANCE (Fanconi anemia complementation group E)

Cases 11 and 12, a sibling pair with common variable immune deficiency (CVID) and acute onset of neuropsychiatric symptoms, both carry an ultrarare missense mutation in *FANCE*. Case 11 met the diagnostic criteria for PANS, characterized by a relapsing-remitting course of abrupt onset OCD, anxiety, and rage. Episodes were commonly coincident with group A streptococcal infection and other infections and always improved after treating the infection followed by a corticosteroid burst. IVIg (aimed to treat the underlying CVID) was associated with a reduced frequency of relapses but did not reduce the severity of relapses. Case 12 was diagnosed with ASD as a child after abrupt loss of speech and eye contact following an infection. The patient regained eye contact and sociability, but apraxia of speech continued. He regressed again later in childhood and soon thereafter was diagnosed with PANS, which was characterized by abrupt onset of OCD, anxiety, aggression (self-injurious behavior and property destruction), and tics. Symptoms improved after treating the infection followed by corticosteroids. The patient subsequently developed acute onset dysarthria and mutism and further evaluation revealed CSF pleocytosis, intracranial hypertension, and GAD antibodies (three times the upper limit of normal). Subsequent relapses responded to addressing infection followed by a corticosteroid burst. Relapse frequency (but not severity) was reduced following the introduction of IVIg to treat CVID. Due to a chronic progressive course, Rituximab was added, which was followed by documented improvements by the speech pathologist that were lost when B cells were repopulated (the speech pathologist was blinded to treatments and B cell repopulation). Consequently, Rituximab was scheduled quarterly. The clinical trajectory improved but deteriorations coincident with infections occurred, so Leflunomide was added as adjunct therapy which has led to further stabilization for 2 years. There is a positive history of inflammatory skin disease on both sides of the family, and a family history of immunodeficiency syndrome, arthritis, lupus-related autoantibodies, recurrent sepsis, and recurrent septic joints.

Both cases carry an ultrarare variant of unknown significance in *VPS13B*. Case 11 carries a maternally transmitted *NOD2* variant that has been described as a risk allele in Crohn’s disease^89, 90^. Case 12 carries another ultrarare, pathogenic, novel variant involved in the DNA repair gene, *THRAP3*, which codes for thyroid hormone receptor-associated protein 3, an RNA processing factor involved in ATR kinase-dependent DDR and the expression of several DNA repair proteins, including FANCL ^91, 92^.

### FANCI (Fanconi anemia, complementation group I)

Case 13 is a neurotypical child who developed a sudden onset of OCD, fatigue, restricted eating, severe anxiety, enuresis, and insomnia following an upper respiratory infection.

Autoimmune thyroiditis was diagnosed based on having high titers of anti-thyroid peroxidase and anti-thyroglobulin antibodies, an elevated TSH, and an ultrasound. A nonsense mutation in *FANCI* was identified. There was a partial response to intravenous glucocorticoids, IVIg, and Rituximab. Case 11 also carries ultrarare variants in two other genes that affect DNA repair.

One is a non-synonymous variant in another FC gene, *FANCP/SLX4*, which may be a benign variant. However, cases 14 and 15 also have an ultrarare variant in this gene (see below). The other is a variant in the DNA repair gene, *LIG4,* which codes for DNA Ligase IV, a key enzyme involved in repairing DNA double-strand breaks through non-homologous end joining and sealing DNA breaks that occur during V(D)J recombination^93, 94^. *LIG4* mutations cause autoimmune and immunodeficiency disorders^94, 95^.

### FANCP/SLX4 Fanconi anemia, complementation group P/Structure-Specific endonuclease subunit)

Cases 14 and 15 are siblings diagnosed with CVID who carry a pathogenic mutation in *TNFRSF13B*, which is one of the more common variants found in immune deficiencies^96, 97^. The same variant has been found in several other PANS cases (unpublished observations). They also share an ultrarare variant in *FANCP/SLX4*, but the relatively low CADD score suggests that the gene alone is not sufficient to explain the severe clinical presentation. Case 14 has a history of acute neuropsychiatric decompensation consistent with PANS, who initially responded to treatment with corticosteroid bursts and IVIg and further stabilized on Anakinra. Case 15 initially presented with anxiety but has regressed substantially, developing a seizure disorder and autoimmune encephalitis (AE) that has partially responded to Rituximab, glucocorticoids, and IVIg. Anakinra has not been effective. MRI indicates atrophy. The siblings also have an ultrarare missense mutation adjacent to a splice acceptor site in *DOCK8*, which has been implicated in immune deficiency and autoimmune disorders^64, 98^. Both also share a variant in *MEFV* (pyrin), an inflammasome regulator gene that is commonly mutated in autoinflammatory disorders^99–102^. Finally, they both have an ultrarare variant in *RMRP*, a noncoding RNA that functions as the RNA component of mitochondrial RNA processing endoribonuclease. This is a very intriguing candidate according to our DNA repair model as it acts as an inhibitor of p53^103, 104^. A comparison between wild-type and mutant alleles using RNAfold shows differences in the secondary structure, suggesting that it could be functional (**Supplementary Figure**).

### FANCA Fanconi Anemia, Complementation Group A

Case 16 is a previously healthy and neurotypical child who presented with abrupt-onset OCD, eating restriction, severe irritability, hyperactivity, aggression, mood swings, sensory amplification, and headaches. Physical exam showed truncal instability, subtle choreiform movements, folliculitis, and findings consistent with enthesitis and arthritis (confirmed on joint ultrasound). Labs indicate persistently elevated vasculitis markers (high vWF Ag and D-dimers after COVID-19), persistently elevated C4, and persistent microcytic anemia thought to be secondary to chronic inflammation. The patient has had a secondary progressive course with relapses coincident with infections (influenza, mycoplasma, COVID-19, and sinusitis) with partial improvement after bacterial infections were treated. Psychotropics and CBT have been minimally helpful. The patient did not tolerate oral or IV steroids, IVIg, or immunomodulation aimed at treating arthritis and systemic inflammation (e.g., methotrexate, sulfasalazine, and azathioprine). There has been a partial response to NSAIDs, and more recently, a solid trajectory of improvement since introducing colchicine. An ultrarare pathogenic variant (according to AlphaMissense) was found in *FANCA*. In addition, ultrarare variants were found in five other genes that have effects on innate immune function, but no direct effect on DNA repair. The most important is a pathogenic variant in *UNC93B1*, a regulator of nucleic acid-sensing toll-like receptors (e.g., TLR3, TLR7, and TLR8) and interferon and STING signaling, suggesting a synergistic interaction with FANCA-related DNA repair defects^105–107^. The others are ultrarare variants of unknown significance in *NLRP1, TICAM1, TYK2*, and *TCN2*. NLRP1, TICAM1, and TYK2 have regulatory effects on the innate immune system, including nucleic acid sensing, and have been implicated in autoimmune disorders^108–111^.

First-degree relatives have migraines, recurrent sinusitis, recurrent pancreatitis, and spondylarthritis. First-degree family members also have anxiety, OCD, ADHD, and depression.

### FANCC Fanconi Anemia, Complementation Group C

Case 17 had a typically developing childhood but developed OCD, ADHD, anxiety symptoms in pre-school, followed by three significant deteriorations over the next couple of years resulting in massive escalation of psychiatric symptoms - OCD, eating restriction, anxiety, irritability, extreme levels of oppositionality, poor sleep, polyuria, school refusal, and refusal to engage in basic routine personal care. Each deterioration resulted in prolonged absences from school and uncontrollable behavior when able to attend. Treatment with an SSRI increased rage and resulted in the emergence of suicidality, which led to the addition of a mood stabilizer.

Eventually, he was diagnosed with ASD. A medical work-up revealed: low C3, low C4, anti-histone antibodies, microcytic anemia, livedo reticularis, enthesitis related arthritis, mild proteinuria, sinusitis, and mild hypothyroidism. Treatment of suspected sinusitis followed by high dose IVIg and a steroid burst resulted in a clear reduction in all psychiatric symptoms, the first improvement seen in two years. Based on the arthritis diagnosis, and concern for relapse after being weaned from steroids, the patient was started on standard of care for arthritis (NSAIDS, methotrexate, sulfasalazine) and was able to return to school. Ongoing ADHD, OCD, and anxiety, which have been relatively mild compared to previous levels, have been controlled with pharmacological and behavioral therapy. There have been minor flares in psychiatric symptoms and enthesitis after significant illnesses (e.g., flu, sinusitis) which are treated with standard-of-care antibiotics for bacterial infections and early introduction of NSAIDs according to PANS treatment guidelines. He has had no further major relapses and has become a good student who is socially engaged at school and home. There is a history of acute rheumatic fever, spondylarthritis, enthesitis related arthritis, psoriasis, and mood disorders in first degree relatives. A pathogenic frameshift variant was found in *FANCC.* FANCC deficiency has been found to mediate microglial pyroptosis and neuronal apoptosis in a mouse spinal cord contusion model, and regulate mitophagy by interacting with Parkin^112, 113^. Interestingly, we have identified several PANS and regressive ASD cases with *PRKN* copy variants (unpublished observations). Ultrarare variants were also found in *PARN*, and *IGLL1,* neither of which has significant direct effects on DNA repair pathways, although *PARN*, which codes for Poly(A)-Specific Ribonuclease, regulates mRNA 3’ end cleavage reactions under DNA-damaging conditions^114^. IGLL1 is a regulator of B cell differentiation and deficiency leads to autosomal recessive agammaglobulinemia^115, 116^. Both variants were scored as benign according to AM, but the CADD score for the *PARN* variant was 26.8 (**Supplementary Table**).

## Discussion

Sixteen cases were identified with ultrarare variants in protein-coding genes related to p53 and FC DNA repair pathways*: PPM1D, ATM, ATR, 53BP1, FANCE, FANCI, FANCP/SLX4, FANCA,* and *FANCC*. These were often accompanied by additional rare variants related to DNA repair and/or immune deficiencies. Most individuals in this study have an underlying neurodevelopmental disorder, with approximately half diagnosed with ASD. The unifying feature for these cases is the distinct, acute onset change in behavior, severe psychiatric symptoms, and loss of function in association with infections and other stressors. Many cases have symptoms consistent with PANS, while others had acute, severe neuropsychiatric decompensation (e.g., mutism, behavioral regression, loss of previously learned skills, cognitive dysfunction) that did not meet clinical criteria for PANS. Co-morbid inflammatory conditions were common, including vasculitis signs (n=2), enthesitis and/or arthritis (n=4), thyroiditis, and/or elevated thyroid antibodies (n=3). Inflammatory conditions were also common among first-degree family members. Three cases had abnormal MRI scans (cases 5, 9, and 15), and one (case 3) had a normal study. Pleocytosis and moderately elevated anti-GAD antibodies were found in case 12. Clinical improvement was observed in most cases following the resolution of infections and the application of immunomodulation (NSAIDS, corticosteroids, IVIg, Rituximab, colchicine, etc.). The findings are congruent with our previous work and provide further support for our hypothesis that DNA repair deficits are a vulnerability factor for neuropsychiatric decompensation.

The DNA repair hypothesis first emerged from our original genetic analysis in which three out of the five immune genes we identified have strong connections to this process: *PPM1D, CHK2*, and *RAG1*^1^. The idea that DNA repair deficits underlie PANS and acute decompensation in NDD is novel, although studies linking DDR to behavior have been described. For example, postmitotic genome instability in neurons can lead to behavioral alterations and neurodegenerative disorders, and genetic studies show that DNA repair genes are involved in genetic subgroups of ASD and NDDs including most of the genes described in this report (e.g., *PPM1D, ATM, ATR, 53BP1, FANCE, FANCI,* and *FANCP/SLX4*)^117–124^.

The two major gene families affecting DNA repair described in this paper code for proteins involved in the p53 and FC pathways. So far, between the current study and our previous report, we have found ultrarare variants in five protein-coding genes that directly affect p53: *PPM1D, ATM*, *CHK2*, *ATR*, and *53BP1*. Among these genes, pathogenic mutations were found in *PPM1D, ATM*, and, in our previous study, *CHK2.* The ultrarare *ATR* and *53BP1* variants described in this paper, however, are variants of unknown significance that will require replication and functional validation. Remarkably, we also found an ultrarare variant in the long non-coding RNA gene, *RMRP*, a p53 inhibitor^103, 104^. *RMRP* expression is induced by immune activators and is increased in autoimmune/autoinflammatory disorders^125^. As described in the results section, structural differences between the mutant and wild-type alleles were predicted by RNAfold, but again, functional validation would be needed to determine whether the *RMRP* variant we identified affects p53 function and DNA repair.

Ultrarare variants were found in five different FC genes: *FANCE, FANCP*/*SLX4*, *FANCI, FANCA,* and *FANCC.* Among these, all are predicted to be pathogenic, except the *FANCP/SLX4* variants. However, the fact that two different ultrarare variants were found in this gene in three cases from two families increases the likelihood they are playing a role. Interestingly, FC proteins interact with components of the p53 DDR pathway, including ATM and ATR (**Figure 2**)^126–129^.

Some researchers have suggested that autoimmune and/or autoinflammatory processes underlie the development of behavioral regression in PANS and in a subgroup of individuals with ASD^34, 130–132^. Impaired DDR may be one mechanism for these associations as it can lead to leakage of nuclear DNA into the cytosol, which can trigger the cGAS-STING pathway, a key component of the type I interferon (IFN-I) anti-viral innate immune response, especially following an infectious disease trigger^65, 133–135^. Mitochondrial DNA breaks and oxidized mitochondrial DNA released into the cytosol can also trigger cGAS-STING^136–138^. AIM2 inflammasome signaling, which leads to the induction of IL-1β and IL-18, is also triggered by cytosolic DNA^139, 140^.

Physiological activation of cGAS-STING is central for the response to foreign viral DNA, and perhaps viral RNA as well^40^. However, studies show that over-activation can lead to autoimmune, autoinflammatory, and neurodegenerative disorders^141–144^. For example, increased IFN-I responses mediated by cGAS-STING due to abnormal clearance of cytosolic DNA and RNA have been implicated in the pathogenesis of autoimmune disorders including SLE and aggressive rheumatoid arthritis^145–148^. Circulating type I IFN levels are elevated in approximately 50% of patients with SLE and IFN receptor inhibition is an approved therapy^149, 150^. A substantial fraction of SLE patients have neuropsychiatric symptoms^151, 152^. It has also been proposed that some neurodegenerative disorders involve cGAS-STING signaling, including Alzheimer’s Disease (AD), Parkinson’s Disease (PD), Amyotrophic Lateral Sclerosis (ALS), Huntington’s disease (HD), and multiple sclerosis (MS)^153–155^. Markers for neurodegenerative disorders are elevated in 27% of patients with psychiatric disease preselected for suspected immunological involvement and phenotypes including catatonia, agitation, and acute onset of symptoms similar to the cases described here, although the genetic data is not available (https://doi.org/10.21203/rs.3.rs-3491787/v1). Early immunological interventions may mitigate some damage, but this needs to be confirmed in clinical trials. However, other forms of damage may accumulate over time that are not responsive to immunological interventions.

In addition to nuclear DNA, disruption of mitochondrial integrity can also lead to the release of mitochondrial DNA into the cytoplasm, which is a potent cGAS-STING trigger^136–138^. A few of the genes described in this report have effects on mitochondria (e.g., *MTO1, SLC13A5, RMRP*). Some FC proteins also regulate mitochondrial DNA replication fork stability that can activate the mtDNA-dependent cGAS/STING response when dysfunctional, and FANCI has been found to regulate PRKN-mediated mitophagy^156, 157^.

A large number of cases had more than one DNA repair gene mutation. This could be due to ascertainment bias since most of the cases we studied had severe symptoms and accompanying neurodevelopmental problems, which could have prompted genetic testing by their physicians. Gene panels rather than WGS per se were performed in many cases, which could cause ascertainment bias due to the selection of genes represented on the commercially available gene panels (**Table**). However, the histories of autoimmune disorders from both parents in many cases suggest true bilineal inheritance of risk variants.

The current paper explores a genetic subgroup linked to impaired DNA repair with acute onset of severe psychiatric symptoms or regression in children associated with infections. It is important to note that non-infectious cellular stressors can also induce DNA damage and activate cGAS-STING, such as cellular injury, heat shock, and oxidative stress^158, 159^. In addition, recent studies suggest that faulty DNA repair could disrupt several cellular processes beyond an innate immune response, such as autophagy, senescence, and apoptosis^160–162^.

Clinically, if our findings are confirmed, they could lead to specific diagnostic tools and new treatments for infection-associated psychiatric episodes and behavioral decompensation in PANS, ASD, and other NDDs in those with DNA repair mutations, targeting putative over-stimulated immune pathways (e.g., type I interferons and AIM2 inflammasomes). To validate our findings, more studies are needed to integrate longitudinal clinical data, genetics, and immunophenotypes. Functional studies on immune cells from patients, and microglia from animal models and induced pluripotent stem cells (iPSCs) are also needed. Studies using iPSC-derived microglia with genetic variants in DNA repair genes are ongoing.

## Acknowledgments

H.M.L. is supported by the National Institute of Child Health and Human Development NIH/NICHD; P30 HD071593 to the Albert Einstein College of Medicine’s Rose F. Kennedy Intellectual and Developmental Disabilities Research Center, and the National Institute of Mental Health, R21MH131740. The Lachman lab also receives support from the Janice C. Blanchard Family Fund and the iPS Cell Research for Ryan Stearn Fund. This work was funded by grants to the J.L.C Swedish Research Council (Grant No. 2019-06082). P.J. van der Spek is supported by EU H2020 grants, an ImmunAID grant (ID: 7792950), and a MOODSTRATIFICATION grant (ID: 754740). The Bioinformatics infrastructure and team are supported by grants from KWF, NWO/ZonMW, and the Dutch Heart Foundation through the BDVA-initiated H2020 Bigmedilytics program on Personalized Medicine. Janet Cunningham is a Gullstrand Fellow at Uppsala University Hospital. Jennifer Frankovich’s research is supported by several foundations including the Neuroimmune Foundation, Lucile Packard Foundation for Children’s Health, the Dollinger Biomarker Core, Stanford Spark, and collaborations on NIH grants. Funders had no role in study design, data collection and analysis, decision to publish, or preparation of the manuscript. We thank the Albert Einstein College of Medicine Epigenomics Shared Core Facility RRID: SCR_023284 for Library preparation, QC, and sequencing, and Yongmei Zhao for technical assistance with the library preps. The authors want to thank the participating families.

## Author contributions

J.L.C. co-wrote the paper, and was involved in the study concept and design; J.F. contributed cases, provided genetic data for cases 16, and 17, and co-wrote the manuscript); R,A.B. carried out WES on case 10); E.P. processed DNA, validated variants, edited manuscript, created figures 1,3; R.F.A. validated variants; N.C.S. contributed case 16; S.B.M. made the WES library for case 10; H.D. involved in study concept; J.M. validated variants); J.E. validated variants; J.M.S. validated variants carried out Alphamissense analysis; G.L. contributed case 1, edited manuscript); S.M.A.S. bioinformatics of variants, created Figure 4; S.S. edited manuscript;P.J.S. study concept and design, bioinformatics; H.M.L.co-wrote paper, study concept and design, analyzed variants, confirmed diagnoses, supervised study. All authors reviewed the manuscript.

## Data availability statement

Variants reported in this paper will be submitted to ClinVar. WES data on case 10 will be made available at GenBank ®

## Supplementary Data

**Supplementary Methods.** An expanded description of the methods and primers used for Sanger DNA sequencing.

**Supplementary Table**. Description of additional gene variants found upon whole exome sequencing or the analysis of gene panels.

**Supplementary Figure. RMRP secondary structures.** The RNAfold web server was used to predict secondary structures for wild-type and mutant (c. C126T) RMRP RNAs. Differences between the two, especially in the centroid secondary structures, are seen.

