## supplemental methods for "Ultrarare Variants in DNA Damage Repair Genes in Pediatric Acute-Onset Neuropsychiatric Syndrome or Acute Behavioral Regression in Neurodevelopmental Disorders"

**Sample preparation and next generation sequencing: WES and WGS**

Genomic DNA (200 ng) extracted from saliva was used. DNA quantity was determined by Qubit (Invitrogen: Thermo Fisher Scientific) and assessed for quality and fragment size through the 4150 TapeStation System (Agilent Technologies, Santa Clara, CA). Pre-captured DNA Libraries were prepared following the protocol using Mechanical Fragmentation and Twist Universal Adapter System (Twist Bioscience) 37138343. These libraries were prepared using the Twist Target Enrichment Protocol and pooled into a single pool for hybridization with Twist Exome 2.0 plus Comprehensive Exome spike-in. The preparation and enrichment of libraries were carried out in accordance with the manufacturer’s instructions. Hybrid-selected libraries were sequenced using the IIIumina Nextseq 2000, employing a paired-end × 150 bases (PE × 150) configuration following standard protocols.

**Variant calling and annotation**

Raw sequencing data was QC and analyzed using GSNAP (http://research-pub.gene.com/gmap/) software for mapping to a reference genome. Adapter-trimmed fastq files were aligned to the human genome (hg38) using bwa (version 0.7.17-r1188; mem algorithm; bwa citation: see https://bio-bwa.sourceforge.net). Bam files were sorted, duplicates marked, and hybrid-selection metrics determined using Picard Tools (version: 2.26.10; bwa citation: see https://bio-bwa.sourceforge.net) modules SortSam, MarkDuplicates, and CollectHsMetrics, respectively; duplicates ranged from 0.18 – 0.2 while mean bait coverage of the duplicates marked bam files in exons and adjacent to exons was between 98 – 126. Base Quality Score Recalibration (BQSR) was performed on the duplicates marked bam files using GATK (version: 4.4.0.0; https://gatk.broadinstitute.org/hc/en-us/articles/360035530852) modules BaseRecalibrator /ApplyBQSR and these output files used as input for variant calling within and nearby to exons, using Strelka (version: 2.9.10) 30013048. For functional annotation VCF file entries that passed all filters (PASS in the FILTER column) were annotated with Annovar (version: Date: 2020-06-08) 20601685. All ultra-rare variants (MAF<0.001) in genes involved in immune disorders, DNA repair, and neurodevelopmental disorders were assessed. We also manually evaluated all genes that had deleterious mutations.

**Evaluation of missense variants**

The predicted pathogenicity of each variant was obtained from the ClinVar database (<https://www.ncbi.nlm.nih.gov/clinvar/>). Variants were aslo evaluated by Combined Annotation Dependent Depletion (CADD), a Phred-like score that is a measure of deleteriousness **30371827 30742610 24487276.** A CADD score represents a ranking, not a prediction, and no threshold is defined. Higher scores are more likely to be deleterious. A score of 20 is commonly used as a cutoff, with scores above 20 predicted to be among the 1.0% most deleterious possible substitutions in the human genome **PMID: 3074261.** CADD scores were obtained using a tool developed by the University of Washington, (https://cadd.gs.washington.edu.) and the CADD_VEP plugin https://useast.ensembl.org/info/docs/tools/vep/script/vep_plugins.html

Finally, missense mutations were also evaluated by AlphaMissense, in which amino acid changes are evaluated by an adaptation of AlphaFold to predict missense variant pathogenicity **37733863**. It combines the structural context of the amino acid substitution and evolutionary conservation to provide a pathogenicity score. AlphaMissense predicts the pathogenicity of all possible single amino acid substitutions in nearly all protein-coding genes, providing the capacity to classify 89% of missense variants.

**Network analysis**

Functional analysis was performed within Ingenuity Pathway Analysis (IPA) (QIAGEN Inc.). All candidate genes were imported in IPA to assess the pathways involved. Using the network tool within IPA, a connectivity network was constructed based on the IPA/QIAGEN Knowledge Base. Both direct and indirect relationships were used to construct the biological network.

**RNAfold**

The secondary structure of RMRP, a long, non-coding RNA was predicted using the RNAfold web server, which was established by the Institute for Theoretical Chemistry, University of Vienna *http://rna.tbi.univie.ac.at/cgi-bin/RNAWebSuite/RNAfold.cgi).

**Validation of selected variants**

The variants found in families 1,2, and 3 were validated saliva DNA. This was obtained using a DNA Genotek kit (cat# OGR-500). DNA was extracted using prepIT purifier (DNAgenotek cat# PT-L2P-5) according to the manufacturer’s instructions, except for the inclusion of a phenol/chloroform purification step (Phenol:Chloroform:Isoamyl Alcohol 25:24:1 saturated with 10mMTris pH8.0, 1mM EDTA; followed by the addition of 1ul glycogen, sodium acetate, pH 5.2 to a final concentration of 300 mM, and an equal volume of Isoamyl Alcohol to precipitate DNA). Genomic DNA was amplified using HotStarTaq DNA Polymerase (Qiagen cat# 203203) according to the manufacturer’s instructions using primers that flank the variants of interest (see list below). PCR products were purified using a QIAquick PCR Purification Kit (Qiagen, catalog# 28104) according to the manufacturer’s instructions. DNA and was sequenced by the standard Sanger dideoxy chain termination method using one of the two PCR primers. We also validated the variants found in the sample we sequenced, case 8. We also validated the *PPM1D* mosaicism in case 1 using both saliva and WBC DNA. The primers used are shown below.

CACNA1S

F-CTCATGTCTCAGGGAGCTGG

R-GGCCTACCTAGGACCCATTG

NLXR1

F-GAGATGAGTTGTGAGGCCCC

R-GTTCCGCTGGACTTCACTGA

ATR

F-CATGTGTATATGCTTCCTTCATTCT

R-ACAGATGTAAAAGCAGTTCTTGGA

CENPJ

F-TCATCAGCAGCTTGTTCTAAAAACA

R-CTGAGTGTCCAAAACCTTGCG

DBH

F-CCATCTTCCTGGTCATCCTG

R-TGGGTTAAGGAGGGTGAGTG

CLPB

F-ACCCAGAGCGGGTTAGGG

R-AGGCACAGCCCCCAAAGT

53BP1

F-TGGGCCTTTCAATCAAGG

R-ATTTTCAGGGGGAACTTGCT

MED1

F-CCACCATCACTGTTCCCTTAG

R-CCGGATTAGACAGCAAACCA

KMT2D

F-GGAGGGTGAGTCAACAAAGC

R-ATTCCACACTACCCCACCTG

THRAP3

F-TGTCCAAAGAGGTTCACATTTGC

R-AGTCACACCCCCTTACCTCA

PPM1D

F-GCTCGAGAGAATGTCCAAGG

R-TCTTTCGCTGTGAGGTTGTG

ATM

F-TCAGCGAAGTGGTGTTCTTG

R-TTTGACAATTACCTGATGAAATTAAA

Complement C7

F-TGAGAGCTGATGAAGGTATTGAACA

R-AATGTGTCCCGTACTGGTCG

RAG2

F-AGTCTTTCTCTGTGCAGCGA

R-CTGCCCCACTGGAGTTTTCC

NOD2 c.C1117T, p.Arg373Cys

F-TCTTTGAGCACTGCTGTTGG

R-GAACTCGGTGCGGATGTACT

NOD2 c.C686T, p.Thr229Met

F-CAGTTATGTCAGCGTCCCCC

R-GTCCAGCCATGCCCACAT

RNASEH2B

F-GGTCTGAAGGCCACCTATGA

R-GCATTTACACATAAGCAACTTACTCC

SP110

F-CCATTCAAATGGCAGATTCC

R-TAGAGACAAGGGCCCAAAAG
