## supplementary figure for "Ultrarare Variants in DNA Damage Repair Genes in Pediatric Acute-Onset Neuropsychiatric Syndrome or Acute Behavioral Regression in Neurodevelopmental Disorders"

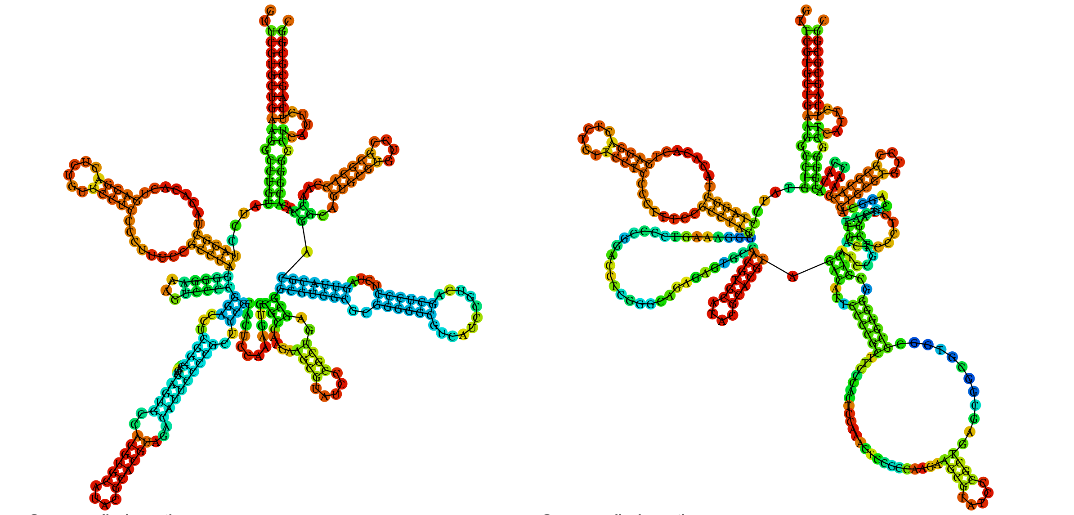

**Optimal secondary structure of wild-type RMRP, minimum free energy -101.30 kcal/mol.**

**Left: MFE secondary structure. Right: centroid secondary structure.**

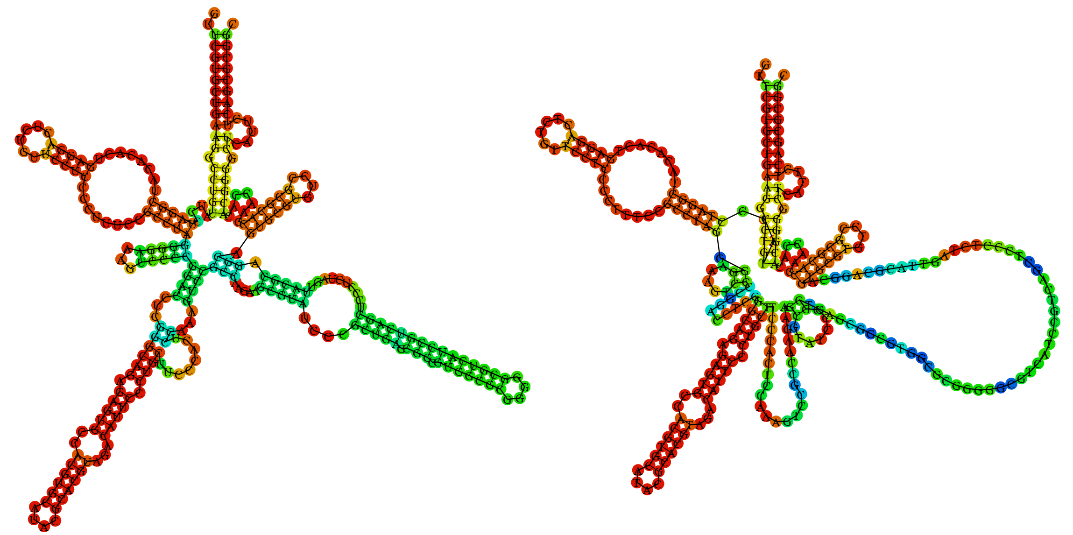

**Optimal secondary structure of mutant RMRP, minimum free energy -105.20 kcal/mol.**

**Left: MFE secondary structure. Right: centroid secondary structure**
